# Postmastectomy Radiotherapy in pN1 Breast Cancer: Survival Outcomes and Prognostic Factors From a Single-Institution Cohort

**DOI:** 10.64898/2026.01.27.26344082

**Authors:** Raksha Madhu Narasimhan, Aren Singh Saini, Kayla Samimi, Ifeanyichukwu Ogobuiro, Xuesen Zhao, Sunwoo Han, Cristiane Takita, Crystal Seldon Taswell

## Abstract

**Purpose/Objectives:** The role of postmastectomy radiotherapy (PMRT) in patients with pathologic N1 (pN1) breast cancer, including triple-negative breast cancer (TNBC), remains controversial in the era of modern systemic therapy. We evaluated the association between PMRT and recurrence-free survival (RFS) and overall survival (OS) and identified prognostic factors in a contemporary single-institution pN1 cohort.

**Materials/Methods:** We retrospectively reviewed female patients with pT1–2N1M0 breast cancer treated with mastectomy between 2016 and 2022. RFS and OS were estimated using Kaplan–Meier methods and compared by PMRT status with log-rank testing. Univariable Cox proportional hazards models assessed associations between clinical factors—including tumor laterality, receptor subtype (TNBC vs non-TNBC), nodal burden, and adjuvant therapies—and survival outcomes, with subgroup analyses by PMRT status and receptor subtype.

**Results:** Fifty-seven patients were included; 22 (38.6%) received PMRT. With a median follow-up of 85 months, PMRT was not associated with improved RFS (median 133 vs 120 months; p=0.256) or OS (not reached vs 195 months; p=0.154). Hormone therapy was significantly associated with improved RFS (HR 0.43; p=0.026) and OS (HR 0.13; p=0.003), while having 2–3 positive lymph nodes predicted worse RFS (HR 2.86; p=0.007). No significant differential benefit from PMRT was observed in patients with TNBC or non-TNBC disease.

**Conclusions:** PMRT was not associated with a survival benefit in this pN1 cohort, including patients with TNBC. Interpretation is limited by modest sample size and statistical power. Outcomes appeared driven by tumor biology, nodal burden, and systemic therapy, supporting individualized PMRT decision-making.

## Introduction

Postmastectomy radiation therapy (PMRT) is a well-established component of adjuvant treatment for high-risk breast cancer, significantly improving locoregional control and survival in patients with extensive nodal involvement [1]. The Danish 82b and 82c studies demonstrated that adding PMRT to mastectomy and systemic therapy markedly reduces locoregional recurrence and improves long-term survival in node-positive disease, firmly establishing PMRT as standard of care for patients with highest risk features [1, 2]. A subsequent meta-analysis by the Early Breast Cancer Trialists’ Collaborative Group (EBCTCG) further confirmed that PMRT confers significant reductions in recurrence and breast cancer mortality, even in patients with limited nodal disease [3].

Despite these data, the use of PMRT in patients with only one to three positive axillary lymph nodes (pN1) remains one of the most debated issues in breast cancer management. Patients with node-negative tumors (pN0) generally do not require PMRT due to very low locoregional recurrence risk, whereas those with four or more involved nodes (N2+) clearly benefit and are routinely treated; the pN1 population lies between these extremes [4]. Notably, the EBCTCG meta-analysis found that PMRT in women with 1–3 positive nodes yielded an absolute ∼8% reduction in 20-year breast cancer mortality [3].

As many patients in those trials lacked contemporary chemotherapy, HER2-targeted therapy, and thorough axillary dissection, it is uncertain whether the same magnitude of benefit applies to today’s pN1 patients [5]. Several tumor and patient factors have been identified that influence locoregional recurrence risk in pN1 disease and thus may modulate the absolute benefit of PMRT. For example, triple-negative or HER2-positive breast cancers and a higher number of positive nodes are associated with greater baseline recurrence risk and potentially greater benefit from PMRT, whereas patients with small, low-grade, hormone receptor–positive tumors and minimal nodal involvement have a lower risk of recurrence [6, 7]. Tumor laterality is another consideration: left-sided cases historically carried higher cardiac toxicity from radiotherapy, though modern techniques have substantially mitigated this risk [8]. Accordingly, the National Comprehensive Cancer Network (NCCN) recommends considering PMRT for patients with 1–3 positive nodes, particularly if high-risk features are present, with omission for node-negative disease [4]. Similarly, a 2016 consensus update by the American Society of Clinical Oncology (ASCO), the American Society for Radiation Oncology (ASTRO), and the Society of Surgical Oncolgoy (SSO) concluded that PMRT significantly lowers recurrence and breast cancer mortality in pN1 overall, but emphasized individualized decision-making, noting that certain low-risk pN1 patients may reasonably be spared from PMRT to avoid undue toxicity [9].

The Selective Use of Postmastectomy Radiotherapy (SUPREMO) trial prospectively evaluated PMRT in intermediate-risk patients by randomizing women with T1–2N1 breast cancer or with high-risk N0 disease to PMRT versus no radiation after mastectomy. Its 10-year results showed no significant improvement in overall survival with PMRT, with approximately 81% 10-year survival in both arms. Although radiotherapy roughly halved the rate of chest wall recurrence, the absolute reduction in locoregional recurrence was under 2%, with no significant impact on distant metastasis or breast cancer mortality [10]. These results underscore that many pN1 patients may not gain a meaningful survival benefit from PMRT in the era of effective hormone therapy, reinforcing the need to better identify which patients truly benefit. This study was therefore undertaken to further clarify the role of PMRT in pN1 breast cancer and to identify prognostic risk factors for survival outcomes in this intermediate-risk population.

## Materials and Methods

We conducted a single-institution retrospective cohort study of female patients diagnosed with pathologic T1N1M0 or T2N1M0 breast cancer who underwent mastectomy between 2016 and 2022. Eligible patients had 1–3 positive axillary lymph nodes on final pathology, no clinical or radiographic evidence of distant metastasis at diagnosis (M0), and documentation of treatment and follow-up. Patients with bilateral breast cancer and prior invasive breast malignancy were excluded.

Clinical and pathological data were abstracted from electronic medical records on November 30, 2023. Researchers had access to identifiable patient information during data collection. Collected variables included age at diagnosis, self-reported race/ethnicity, tumor size and laterality, histology, estrogen receptor (ER) status, progesterone receptor (PR) status, HER2 status, and use of adjuvant therapies including chemotherapy, endocrine therapy, HER2-directed therapy, and receipt of postmastectomy radiation therapy (PMRT). The variables were selected based on clinical relevance and prior literature [11–13].

The primary outcomes were recurrence-free survival (RFS) and overall survival (OS). RFS was defined as elapsed time from diagnosis to recurrence, death from any cause, or last follow-up. OS was defined as elapsed time from diagnosis to death or last follow-up.

Descriptive statistics were used to summarize baseline characteristics. Continuous variables were summarized with mean, median, standard deviation (SD) and interquartile rage (IQR), and two independent groups were compared using Wilcoxon rank-sum test. Categorical variables were tabulated with frequency and percentage, and two groups were compared using Chi-squared test or Fisher’s exact test. Kaplan-Meier method were employed to estimate RFS and OS, and groups were compared using the log-rank test. Cox proportional hazards regression models were used to evaluate associations between prognostic factors and RFS and OS. Analyses were applied for the whole cohort and stratified by receipt of postmastectomy radiotherapy (PMRT): PMRT and non-PMRT cohorts. To explore the effect of PMRT by triple-negative receptor status, we also conducted the corresponding subset analyses. Across all the regression modeling, multivariable analyses were not performed due to the small sample size. Statistical significance was defined as a two-sided p-value <0.05. All analyses were performed using SAS version 9.4.

This study was conducted using data from an institutional breast cancer registry approved by the University of Miami Institutional Review Board (IRB #20090856). The registry includes both retrospectively and prospectively collected clinical data from patients treated for breast cancer within the Department of Radiation Oncology at the University of Miami. For the purposes of this analysis, only retrospectively collected data were used. The data was accessed for research purposes on December 14^th^, 2023. In accordance with the approved protocol, retrospective data collection was conducted under a waiver of informed consent and a waiver of HIPAA authorization. During data abstraction, IRB-approved study personnel had access to identifiable patient information contained within the medical record. All data were subsequently de-identified prior to statistical analysis, and no identifiable information was available to investigators during or after analysis. No direct interaction with patients occurred, and no patients were contacted for this study. Data were abstracted from existing medical records and entered into a secure, HIPAA-compliant REDCap database accessible only to IRB-approved study personnel.

## Results

A total of 57 patients with pT1-2N1M0 breast cancer who underwent mastectomy were included in the study, of whom 22 (38.6%) received postmastectomy radiation therapy (PMRT) and 35 (61.4%) did not. Baseline demographic and clinical characteristics were compared between the two groups, with no statistically significant differences observed in all the variables (p >.05). Of the 54 patients with reported receptor status, the vast majority were hormone receptor–positive and HER2– (83.4% overall; 77.2% without PMRT vs. 94.7% with PMRT), while triple-negative tumors accounted for 16.7% (22.9% without PMRT vs. 5.3% with PMRT). Receptor subtype distribution did not differ significantly between treatment groups (Table 1). For the whole study cohort, median follow-up from diagnosis was 85 months (IQR: 43 to 156).

**Table 1.** Demographics & Clinical Variables.

|  | Total<br>(N=57)<br>N (%) | Non-PMRT<br>(N=35)<br>N (%) | PMRT<br>(N=22)<br>N (%) | P-value |
| --- | --- | --- | --- | --- |
| **Age at Diagnosis** |  |  |  |  |
| Mean (SD) | 57.3 (12.5) | 58.5 (13) | 55.5 (11.7) | 0.3962<sup>1</sup> |
| Median (IQR) | 57 (50, 63) | 57.0 (50, 65) | 56.0 (44, 63) |  |
| Min, Max | 29, 88 | 29, 88 | 37, 75 |  |
| **Race** |  |  |  | 1.0000<sup>2</sup> |
| Black | 8 (14.0) | 5 (14.3) | 3 (13.6) |  |
| White | 49 (86.0) | 30 (85.7) | 19 (86.4) |  |
| **Ethnicity** |  |  |  | 0.9527<sup>3</sup> |
| Non-Hispanic | 36 (63.2) | 22 (62.9) | 14 (63.6) |  |
| Hispanic | 21 (36.8) | 13 (37.1) | 8 (36.4) |  |

**Table 1. Demographics & Clinical Variables**
|  | Total<br>(N=57)<br>N (%) | Non-PMRT<br>(N=35)<br>N (%) | PMRT<br>(N=22)<br>N (%) | P-value |
| --- | --- | --- | --- | --- |
| **Laterality** |  |  |  | 0.3261<sup>2</sup> |
| Right | 27 (47.4) | 14 (40.0) | 13 (59.1) |  |
| Left | 29 (50.9) | 20 (57.1) | 9 (40.9) |  |
| Paired Site | 1 (1.8) | 1 (2.9) | 0 (0.0) |  |
| **Cancer Type, n (%)** | 49 (100) | 32 (100) | 17 (100) | 0.1139<sup>2</sup> |
| IDC | 45 (91.8) | 31 (96.9) | 14 (82.4) |  |
| ILC | 4 (8.2) | 1 (3.1) | 3 (17.6) |  |
| **Chemotherapy, n (%)** | 55 (100) | 34 (100) | 21 (100) | 0.5764<sup>2</sup> |
| None | 20 (36.4) | 14 (41.2) | 6 (28.6) |  |
| NOS | 4 (7.3) | 3 (8.8) | 1 (4.8) |  |
| Single Agent | 3 (5.5) | 1 (2.9) | 2 (9.5) |  |
| Multiple Agents | 28 (50.9) | 16 (47.1) | 12 (57.1) |  |
| **Multiple Chemotherapy, n (%)** | 55 (100) | 34 (100) | 21 (100) | 0.4674<sup>3</sup> |
| None/NOS/Single | 27 (49.1) | 18 (52.9) | 9 (42.9) |  |
| Multiple Agents | 28 (50.9) | 16 (47.1) | 12 (57.1) |  |
| **Hormone Therapy, n (%)** | 56 (100) | 34 (100) | 22 (100) | 0.0815<sup>2</sup> |
| No | 19 (33.9) | 15 (44.1) | 4 (18.2) |  |
| Yes | 37 (66.1) | 19 (55.9) | 18 (81.8) |  |
| **Immunotherapy** |  |  |  | 0.2763<sup>2</sup> |

**Table 1. Demographics & Clinical Variables**
|  | Total<br>(N=57)<br>N (%) | Non-PMRT<br>(N=35)<br>N (%) | PMRT<br>(N=22)<br>N (%) | P-value |
| --- | --- | --- | --- | --- |
| No | 54 (94.7) | 32 (91.4) | 22 (100.0) |  |
| Yes | 3 (5.3) | 3 (8.6) | 0 (0.0) |  |
| **Pathologic Tumor Size, n (%)** | 56 (100) | 35 (100) | 21 (100) | 0.8922<sup>2</sup> |
| p1a | 1 (1.8) | 1 (2.9) | 0 (0.0) |  |
| p1b | 4 (7.1) | 3 (8.6) | 1 (4.8) |  |
| p1c | 16 (28.6) | 9 (25.7) | 7 (33.3) |  |
| p2 | 35 (62.5) | 22 (62.9) | 13 (61.9) |  |
| **Surgical Pathologic Stage** |  |  |  | 0.4209<sup>2</sup> |
| 1a | 2 (3.5) | 1 (2.9) | 1 (4.5) |  |
| 1b | 6 (10.5) | 2 (5.7) | 4 (18.2) |  |
| 1c | 21 (36.8) | 12 (34.3) | 9 (40.9) |  |
| 2a | 26 (45.6) | 18 (51.4) | 8 (36.4) |  |
| 2b | 2 (3.5) | 2 (5.7) | 0 (0.0) |  |
| **Receptor Status, n (%)** | 54 (100) | 35 (100) | 19 (100) | 0.1075<sup>2</sup> |
| Triple Negative | 9 (16.7) | 8 (22.9) | 1 (5.3) |  |
| ER+PR+HER2- | 42 (77.8) | 24 (68.6) | 18 (94.7) |  |
| ER+PR-HER2- | 3 (5.6) | 3 (8.6) | 0 (0.0%) |  |
| **Number of Positive Lymph**<br>**Nodes, n (%)** | 51 (100) | 32 (100) | 19 (100) | 0.3317<sup>2</sup> |

**Table 1. Demographics & Clinical Variables**
|  | Total<br>(N=57)<br>N (%) | Non-PMRT<br>(N=35)<br>N (%) | PMRT<br>(N=22)<br>N (%) | P-value |
| --- | --- | --- | --- | --- |
| 1 | 39 (76.5) | 24 (75.0) | 15 (78.9) |  |
| 2 | 9 (17.6) | 7 (21.9) | 2 (10.5) |  |
| 3 | 3 (5.9) | 1 (3.1) | 2 (10.5) |  |
| **Recurrence Occurred** |  |  |  | 0.424<sup>3</sup> |
| No | 26 (45.6) | 14 (40.0) | 12 (54.5) |  |
| Yes | 31 (54.4) | 21 (60.0) | 10 (45.5) |  |
| Local | 5 (8.8) | 4 (11.4) | 1 (4.5) |  |
| Nodal | 2 (3.5) | 2 (5.7) | 0 (0.0) |  |
| Systemic | 24 (42.1) | 15 (42.9) | 9 (40.9) |  |
| **Vital Status** |  |  |  | 0.153<sup>3</sup> |
| Dead | 12 (21.1) | 10 (28.6) | 2 (9.1) |  |
| Alive | 45 (78.9) | 25 (71.4) | 20 (90.9) |  |
| <sup>1</sup>Equal variance two sample t-test; <sup>2</sup>Fisher Exact p-value; <sup>3</sup>Chi-Square p-value; SD: Standard deviation; IQR: Interquartile Range. |  |  |  |  |

### Recurrence-Free and Overall Survival

Of the 57 patients, there were 31 recurrence events–5 local, 2 nodal, and 24 systemic recurrences –and 12 deaths (Table 1). For the whole cohort, median PFS and OS were 133 months (95% CI: 72–155) and not estimable (NE), respectively (Figure 1). When stratified by receipt of PMRT, there was no statistically significant difference in RFS or OS between the PMRT and non-PMRT groups. Median RFS was 133 months (95% CI: 72–NE) in the PMRT group versus 120 months (95% CI: 33–155) in the non-PMRT group (log-rank p=0.256). Median OS was not reached in the PMRT group and was 195 months (95% CI: 124–NE) in the non-PMRT group (log-rank p=0.154) (Figure 2).

**Figure 1.** Kaplan–Meier estimates of RFS and OS for all patients. Tick marks denote censored observations.

**Figure 2.** Kaplan–Meier estimates of RFS and OS stratified by PMRT status. Tick marks denote censored observations. Survival comparisons were performed using the log-rank test.

### Prognostic Factors of Recurrence-Free Survival

In univariable Cox regression analyses across the entire cohort (n=57), receipt of hormone therapy was significantly associated with improved RFS (HR=0.43, 95% CI: 0.21–0.90, p=0.026), while having two or three positive lymph nodes was associated with worse RFS (HR=2.86, 95% CI: 1.34–6.11, p=0.007). Receptor status was not significantly associated with RFS in the overall cohort (Table 2). Though the difference was not statistically significant, there was a trend toward improved RFS in the PMRT group (HR=0.65, 95% CI: 0.30-1.38, p=0.263) (Figure 2).

**Table 2.**
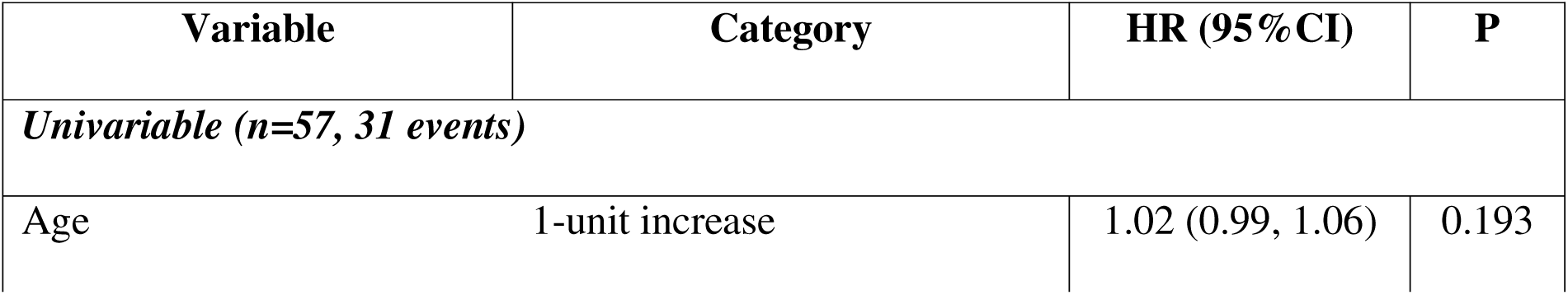
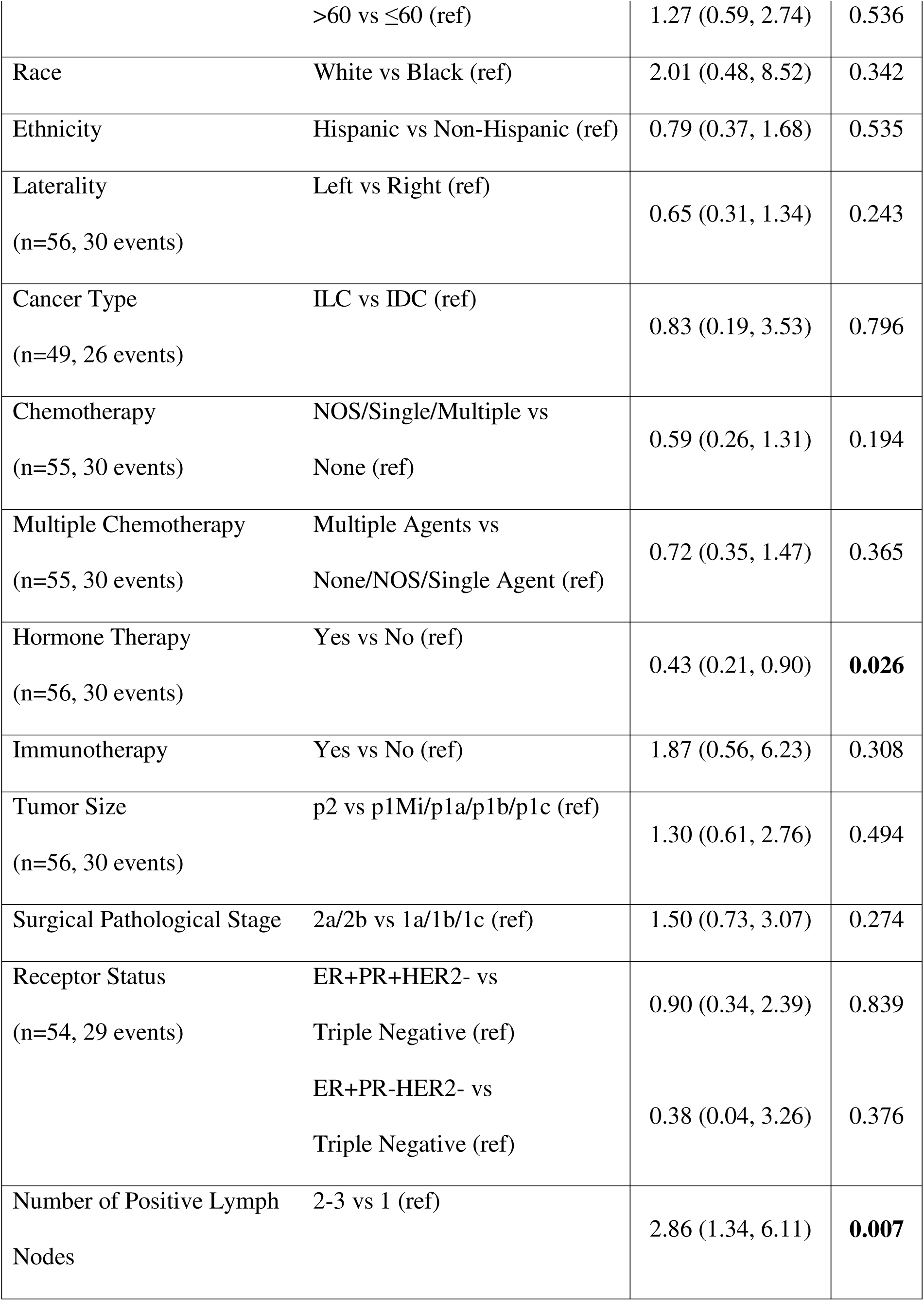
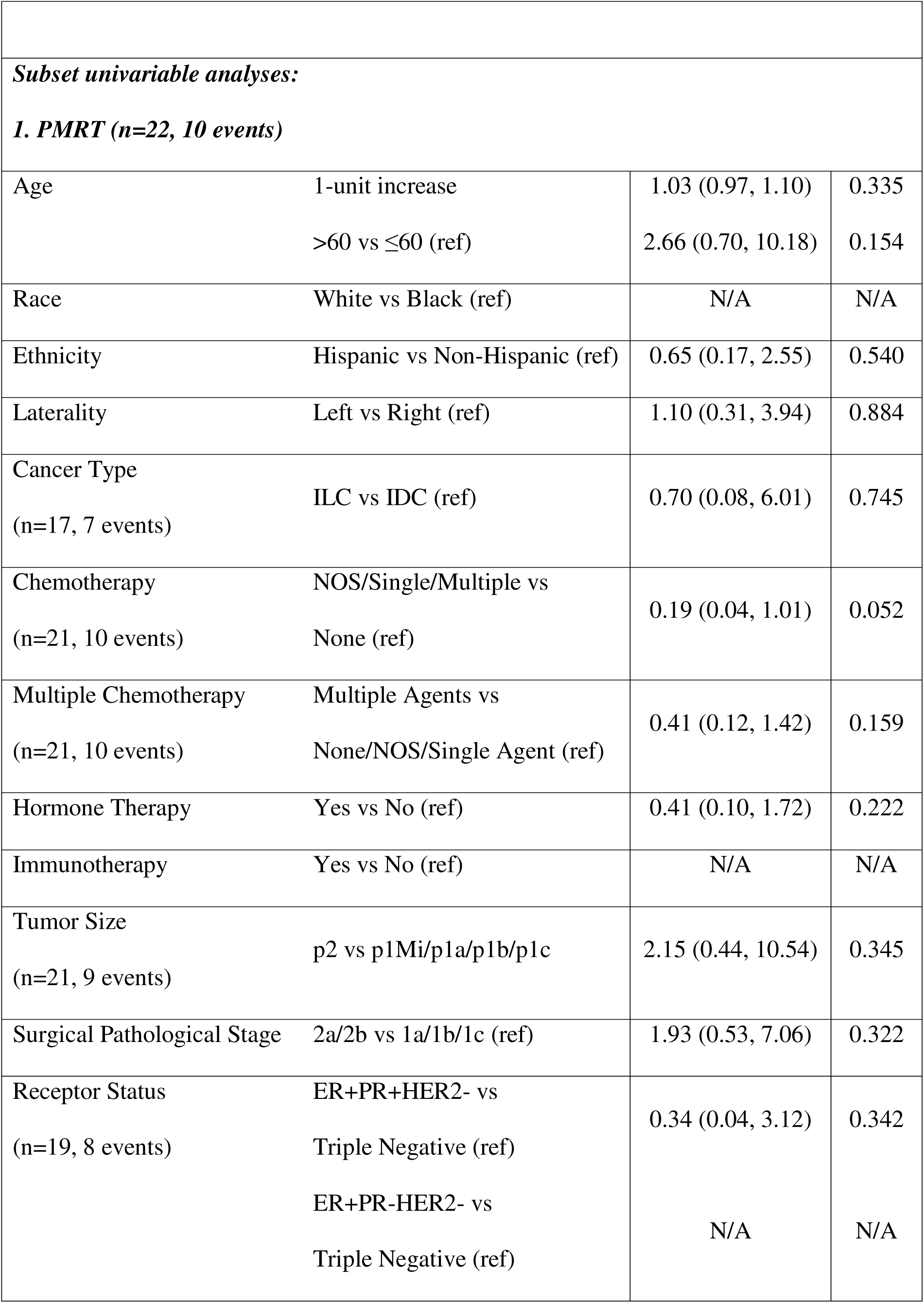
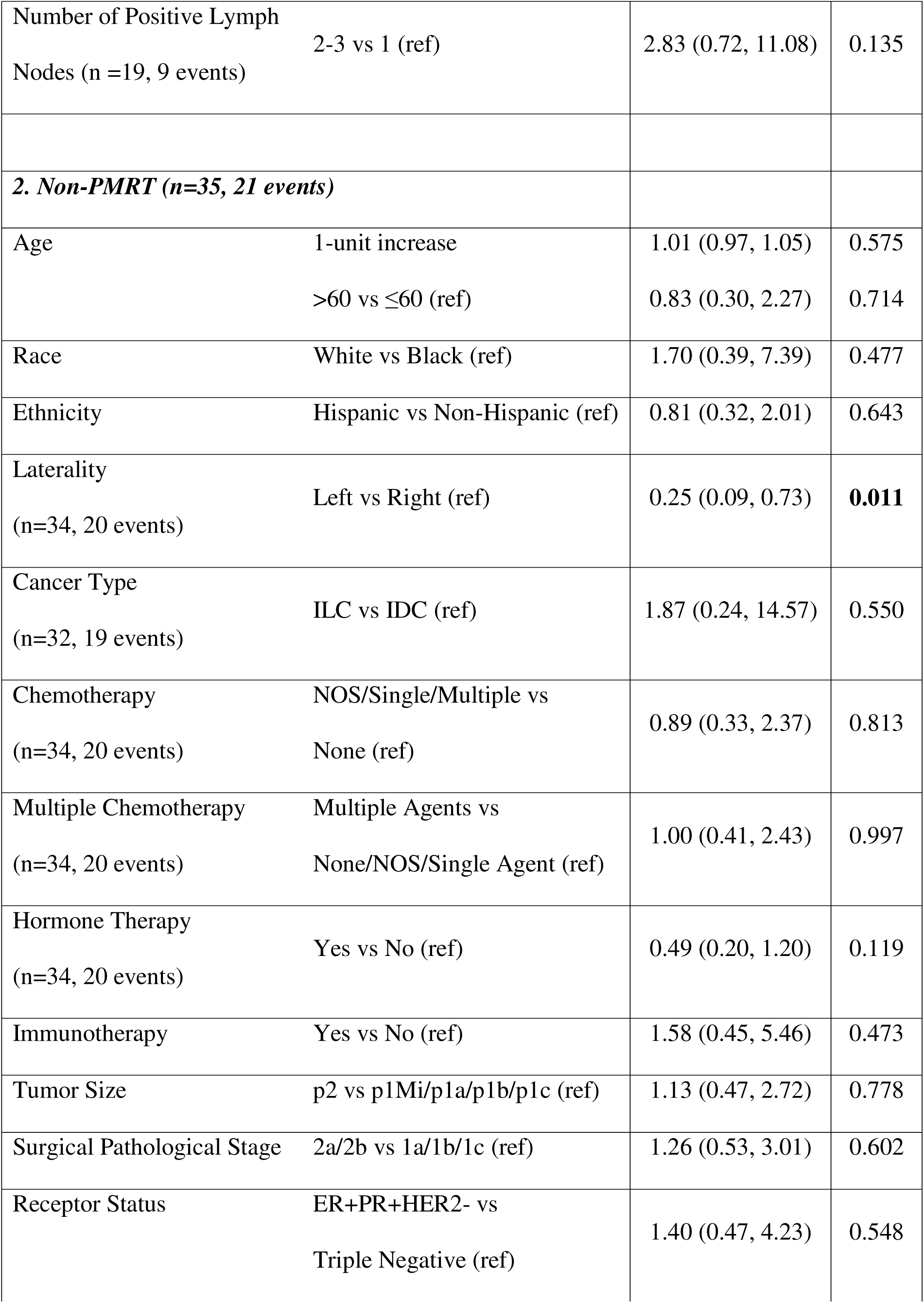
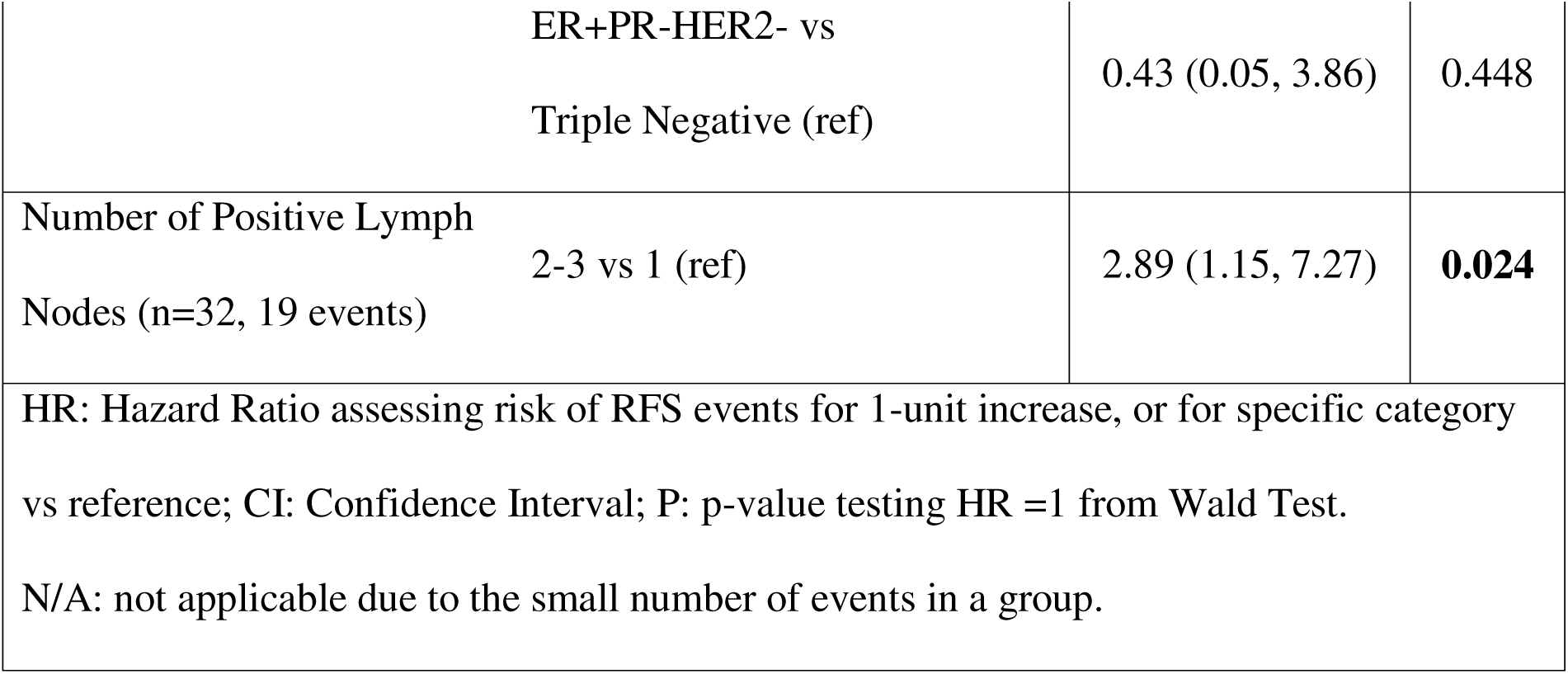
Cox Regression Analyses Assessing Risk Factors for RFS.

In subset analyses with patients who did not receive PMRT (n=35), univariable Cox models identified left-sided tumor laterality as a significant predictor of improved RFS compared to right-sided disease (HR=0.25, 95% CI: 0.09–0.73, p=0.011). A higher nodal burden (2-3 nodes) was also associated with increased recurrence risk compared to single node in this subgroup (HR=2.89, 95% CI: 1.15–7.27, p=0.024). Receptor subtype was not significantly associated with RFS in the non-PMRT subgroup (Table 2). In the subset analyses with PMRT group (n=22), none of prognostic factors was identified as significant predictors.

### Prognostic Factors of Overall Survival

In the analyses of OS for the entire cohort, hormone therapy was significantly associated with improved survival (HR=0.13, 95% CI: 0.04–0.50, p=0.003), and ER+PR+HER2− receptor status was also significantly associated with longer OS relative to triple-negative disease (HR=0.25, 95% CI: 0.08–0.80, p=0.019) (Table 3). There were no statistically significant differences in OS between the PMRT group and non-PMRT group (HR=0.33, 95% CI: 0.07-1.51, p=0.154) (Figure 2).

**Table 3.**
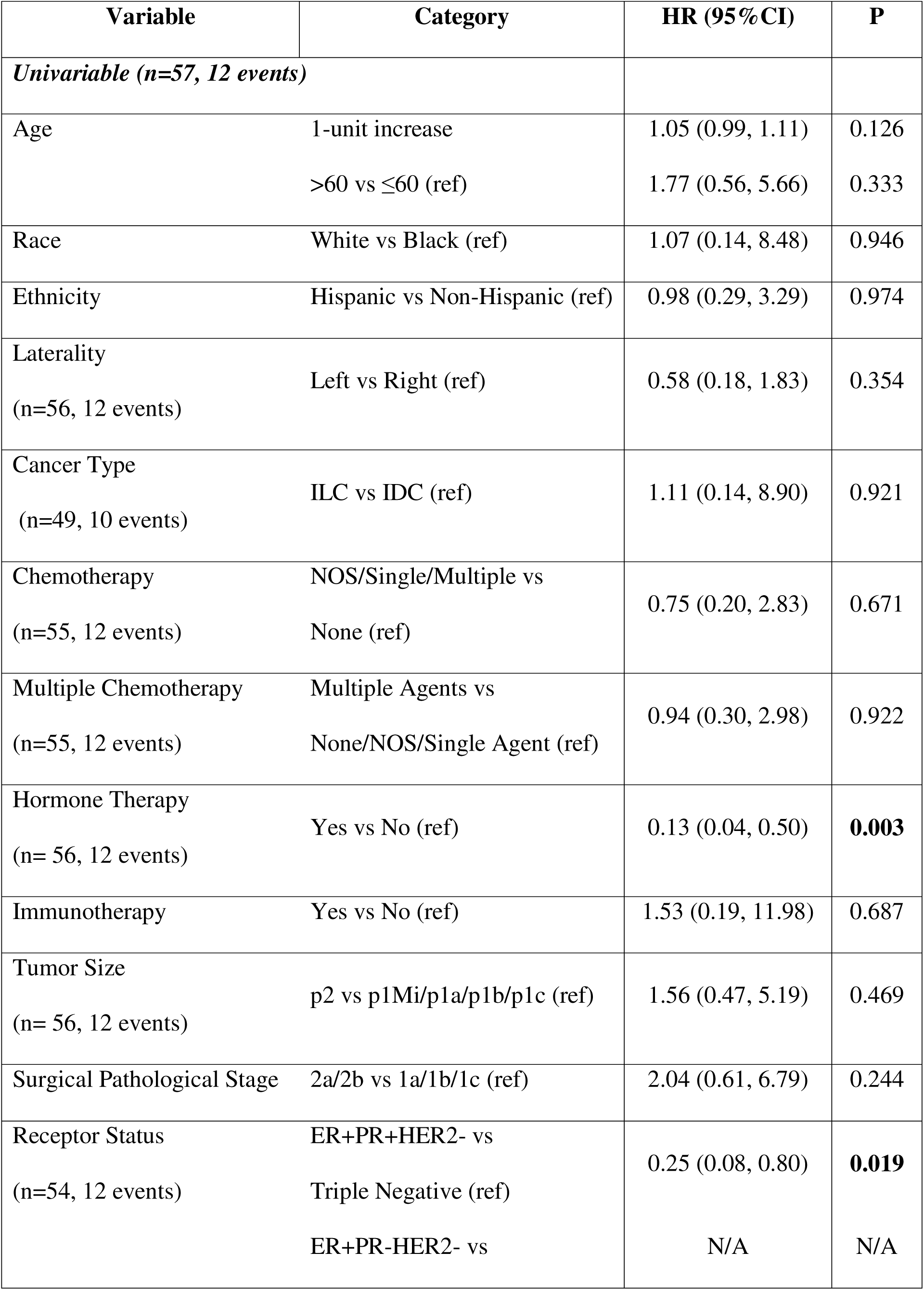
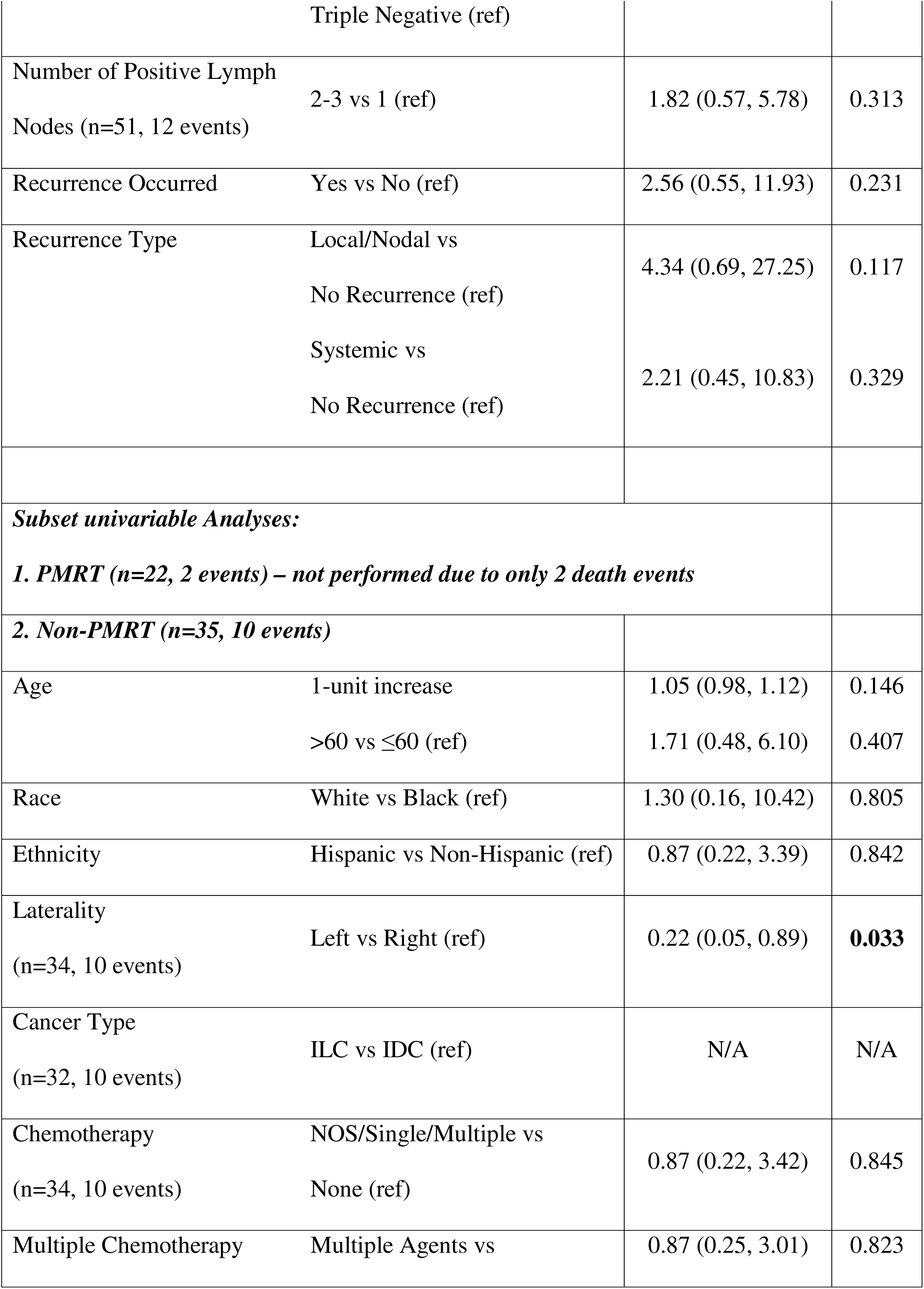
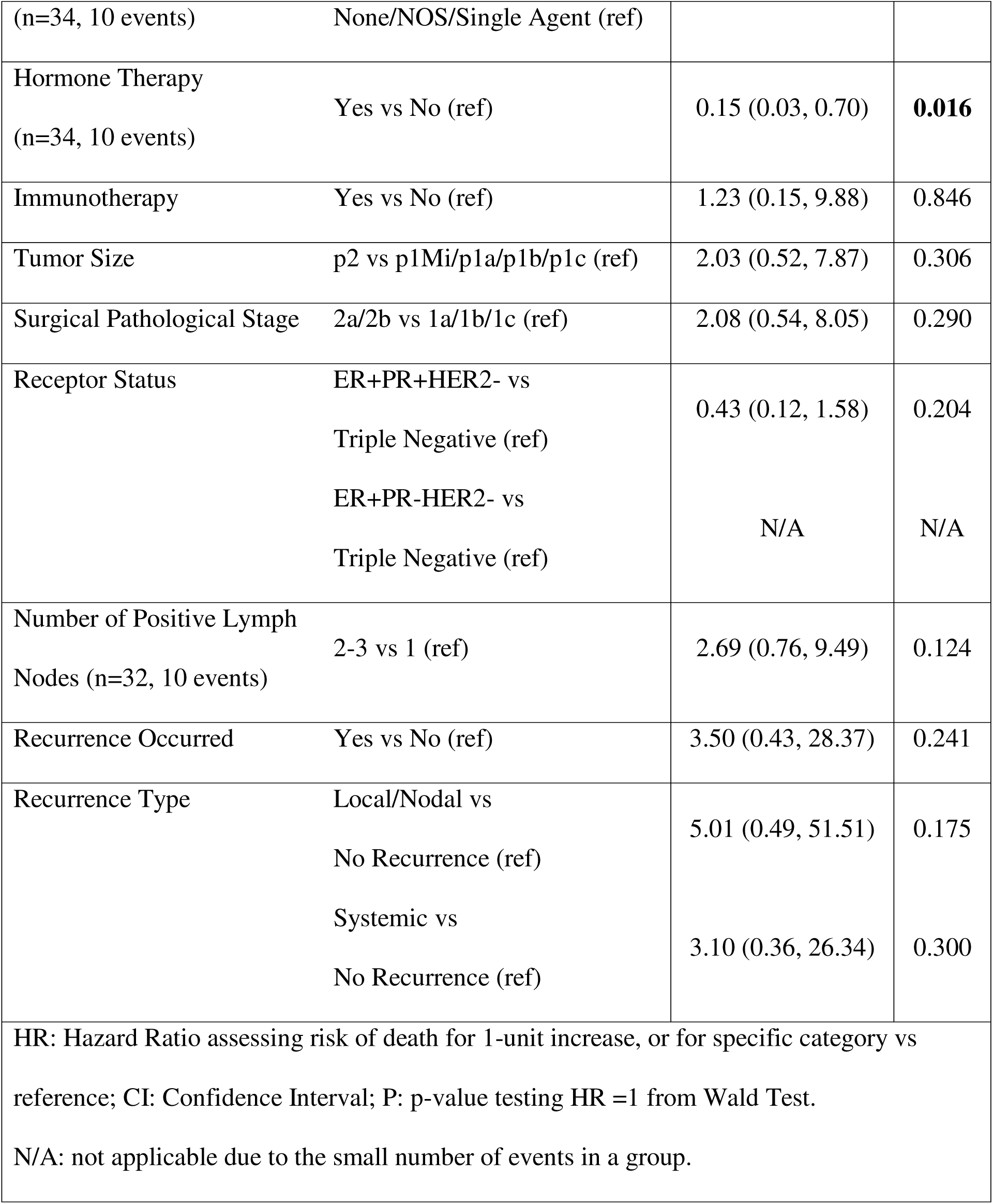
Cox Regression Analyses Assessing Risk Factors for OS.

| Variable | Category | HR (95%CI) | P |
| --- | --- | --- | --- |
| *Univariable (n=57, 12 events)* |  |  |  |
| Age | 1-unit increase | 1.05 (0.99, 1.11) | 0.126 |
|  | >60 vs ≤60 (ref) | 1.77 (0.56, 5.66) | 0.333 |
| Race | White vs Black (ref) | 1.07 (0.14, 8.48) | 0.946 |
| Ethnicity | Hispanic vs Non-Hispanic (ref) | 0.98 (0.29, 3.29) | 0.974 |
| Laterality<br>(n=56, 12 events) | Left vs Right (ref) | 0.58 (0.18, 1.83) | 0.354 |
| Cancer Type<br>(n=49, 10 events) | ILC vs IDC (ref) | 1.11 (0.14, 8.90) | 0.921 |
| Chemotherapy<br>(n=55, 12 events) | NOS/Single/Multiple vs<br>None (ref) | 0.75 (0.20, 2.83) | 0.671 |
| Multiple Chemotherapy<br>(n=55, 12 events) | Multiple Agents vs<br>None/NOS/Single Agent (ref) | 0.94 (0.30, 2.98) | 0.922 |
| Hormone Therapy<br>(n= 56, 12 events) | Yes vs No (ref) | 0.13 (0.04, 0.50) | **0.003** |
| Immunotherapy | Yes vs No (ref) | 1.53 (0.19, 11.98) | 0.687 |
| Tumor Size<br>(n= 56, 12 events) | p2 vs p1Mi/p1a/p1b/p1c (ref) | 1.56 (0.47, 5.19) | 0.469 |
| Surgical Pathological Stage | 2a/2b vs 1a/1b/1c (ref) | 2.04 (0.61, 6.79) | 0.244 |
| Receptor Status<br>(n=54, 12 events) | ER+PR+HER2- vs<br>Triple Negative (ref) | 0.25 (0.08, 0.80) | **0.019** |
|  | ER+PR-HER2- vs | N/A | N/A |
|  | Triple Negative (ref) |  |  |
| Number of Positive Lymph<br>Nodes (n=51, 12 events) | 2-3 vs 1 (ref) | 1.82 (0.57, 5.78) | 0.313 |
| Recurrence Occurred | Yes vs No (ref) | 2.56 (0.55, 11.93) | 0.231 |
| Recurrence Type | Local/Nodal vs<br>No Recurrence (ref) | 4.34 (0.69, 27.25) | 0.117 |
|  | Systemic vs<br>No Recurrence (ref) | 2.21 (0.45, 10.83) | 0.329 |
| *Subset univariable Analyses:* |  |  |  |
| *1. PMRT (n=22, 2 events) – not performed due to only 2 death events* |  |  |  |
| *2. Non-PMRT (n=35, 10 events)* |  |  |  |
| Age | 1-unit increase | 1.05 (0.98, 1.12) | 0.146 |
|  | >60 vs ≤60 (ref) | 1.71 (0.48, 6.10) | 0.407 |
| Race | White vs Black (ref) | 1.30 (0.16, 10.42) | 0.805 |
| Ethnicity | Hispanic vs Non-Hispanic (ref) | 0.87 (0.22, 3.39) | 0.842 |
| Laterality<br>(n=34, 10 events) | Left vs Right (ref) | 0.22 (0.05, 0.89) | **0.033** |
| Cancer Type<br>(n=32, 10 events) | ILC vs IDC (ref) | N/A | N/A |
| Chemotherapy<br>(n=34, 10 events) | NOS/Single/Multiple vs<br>None (ref) | 0.87 (0.22, 3.42) | 0.845 |
| Multiple Chemotherapy | Multiple Agents vs | 0.87 (0.25, 3.01) | 0.823 |
| (n=34, 10 events) | None/NOS/Single Agent (ref) |  |  |
| Hormone Therapy<br>(n=34, 10 events) | Yes vs No (ref) | 0.15 (0.03, 0.70) | **0.016** |
| Immunotherapy | Yes vs No (ref) | 1.23 (0.15, 9.88) | 0.846 |
| Tumor Size | p2 vs p1Mi/p1a/p1b/p1c (ref) | 2.03 (0.52, 7.87) | 0.306 |
| Surgical Pathological Stage | 2a/2b vs 1a/1b/1c (ref) | 2.08 (0.54, 8.05) | 0.290 |
| Receptor Status | ER+PR+HER2- vs<br>Triple Negative (ref) | 0.43 (0.12, 1.58) | 0.204 |
|  | ER+PR-HER2- vs<br>Triple Negative (ref) | N/A | N/A |
| Number of Positive Lymph<br>Nodes (n=32, 10 events) | 2-3 vs 1 (ref) | 2.69 (0.76, 9.49) | 0.124 |
| Recurrence Occurred | Yes vs No (ref) | 3.50 (0.43, 28.37) | 0.241 |
| Recurrence Type | Local/Nodal vs<br>No Recurrence (ref) | 5.01 (0.49, 51.51) | 0.175 |
|  | Systemic vs<br>No Recurrence (ref) | 3.10 (0.36, 26.34) | 0.300 |
| HR: Hazard Ratio assessing risk of death for 1-unit increase, or for specific category vs reference; CI: Confidence Interval; P: p-value testing HR =1 from Wald Test.<br>N/A: not applicable due to the small number of events in a group. |  |  |  |

Among patients who did not receive PMRT (n=35), the subset analyses determined that left-sided tumor laterality (HR=0.22, 95% CI: 0.05–0.89, p=0.033) and receipt of hormone therapy (HR=0.15, 95% CI: 0.03–0.70, p=0.016) were significantly associated with improved OS. Due to the limited number of death events (n=2) in the PMRT group, the corresponding subset analysis was not performed (Table 3).

### Receptor Status Analysis

Kaplan–Meier analyses were performed stratified by receptor subtype as triple-negative versus non–triple-negative (Figure 3, Figure 4). Among patients with triple-negative breast cancer (n=9), no statistically significant difference in RFS or OS was observed between PMRT and non-PMRT groups (log-rank p >.05) (Figure 3). Similarly, among patients with non–triple-negative receptor status (n=45), PMRT was not associated with a statistically significant improvement in RFS or OS (low-rank p >.05) (Figure 4).

**Figure 3.** Kaplan–Meier estimates of RFS and OS stratified by PMRT status among patients with triple-negative breast cancer (N=9). Tick marks denote censored observations. Survival comparisons were performed using the log-rank test.

**Figure 4.** Kaplan–Meier estimates of RFS and OS stratified by PMRT status among patients with non–triple-negative breast cancer (N=45). Tick marks denote censored observations. Survival comparisons were performed using the log-rank test.

In subset analyses of patients who did not have triple negative receptor (n=45), univariable Cox model identified hormone therapy as a significant predictor of improved RFS compared to right-sided disease (HR=0.21, 95% CI: 0.08–0.57, p=0.002). Having 2-3 lymph nodes was also associated with increase recurrence risk compared to single node in this subgroup (HR=2.53, 95% CI: 1.07–5.96, p=0.034). However, no prognostic factors was identified as significant predictors in the subset analyses with triple negative receptor group (n=9) (Supplemental Table 1).

**Supplemental Table 1:**
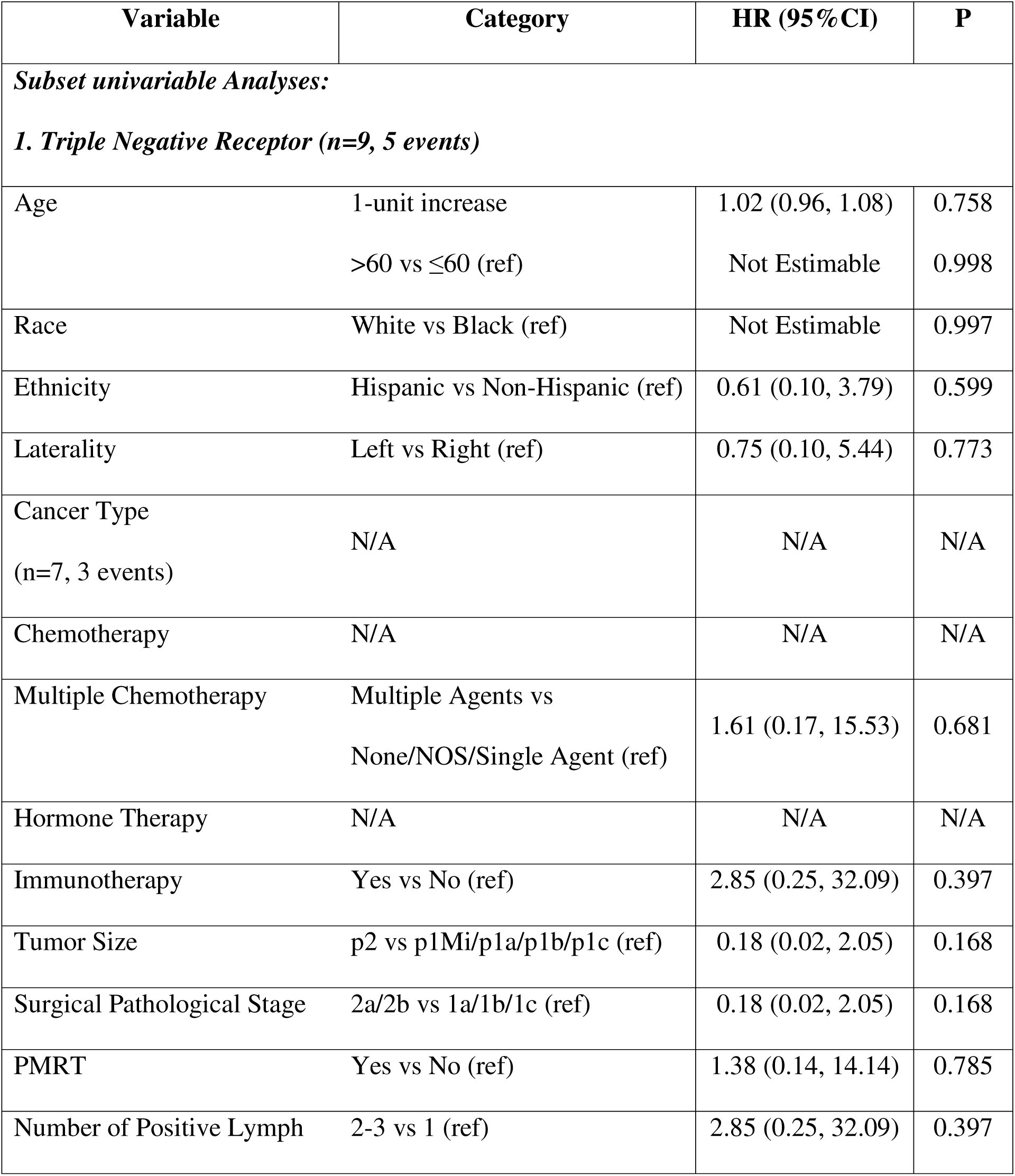
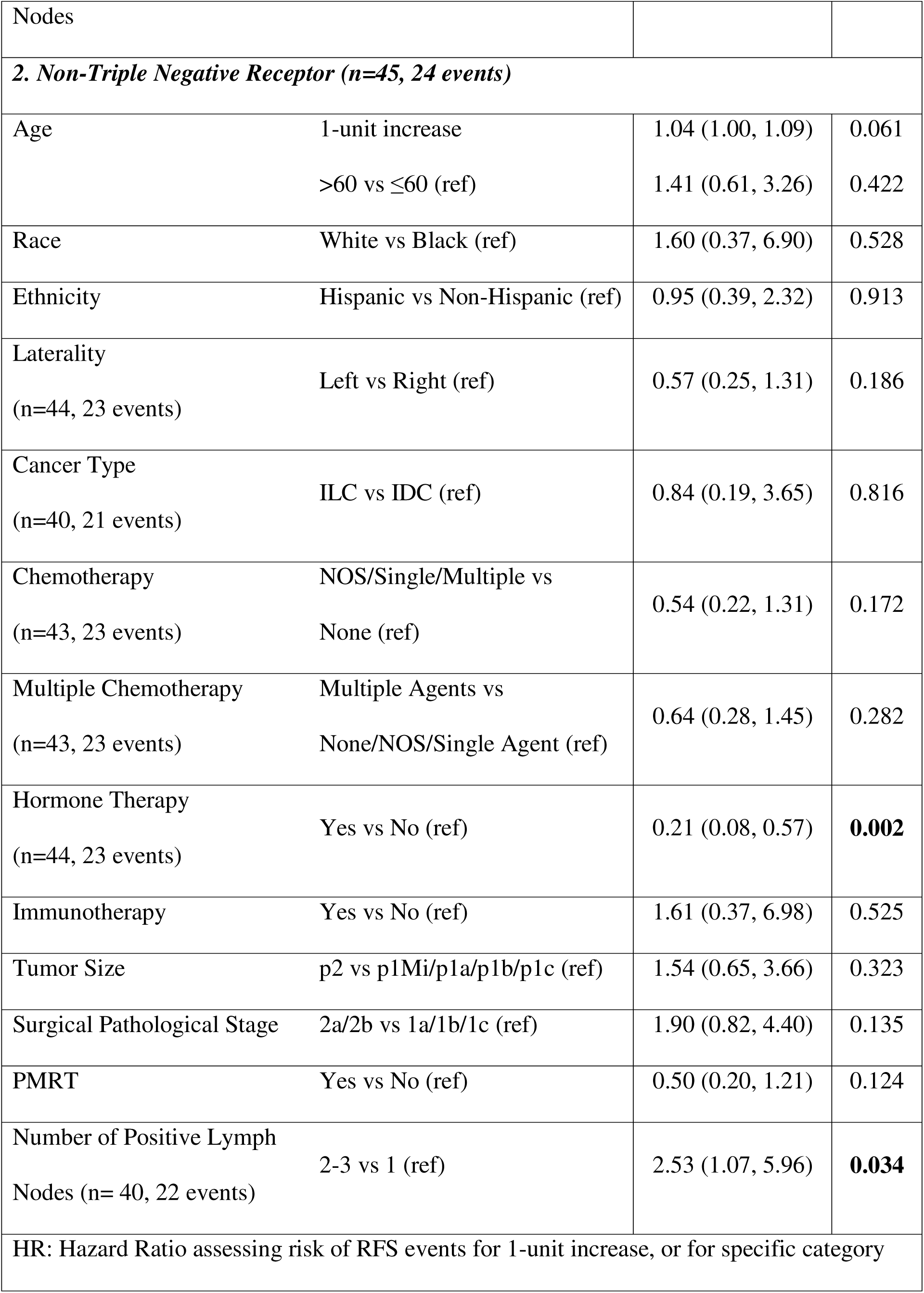
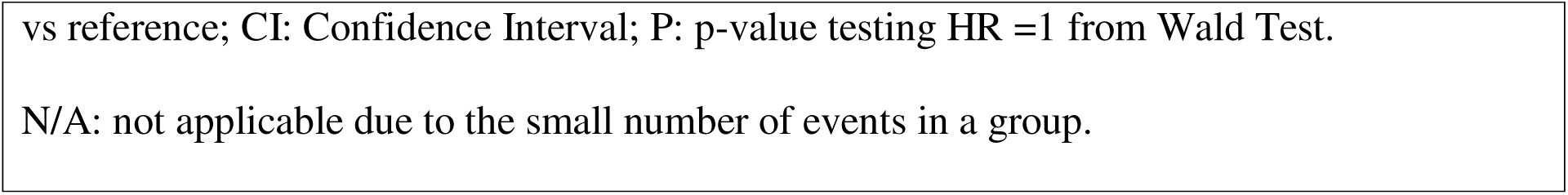
Cox Regression Analyses Assessing Risk Factors for RFS by Receptor Status (Triple negative vs. Non-triple negative)

For patients who did not have triple negative receptor (n=45), age (1-year increase HR=1.14, 95% CI: 1.01–1.29, p=0.039) and hormone therapy (HR=0.12, 95% CI: 0.03–0.56, p=0.007) were identified as significant factors for OS. In patients who have the triple negative receptor type (n=9), no prognostic factors were significantly associated with OS (Supplemental Table 2).

**Supplemental Table 2:**
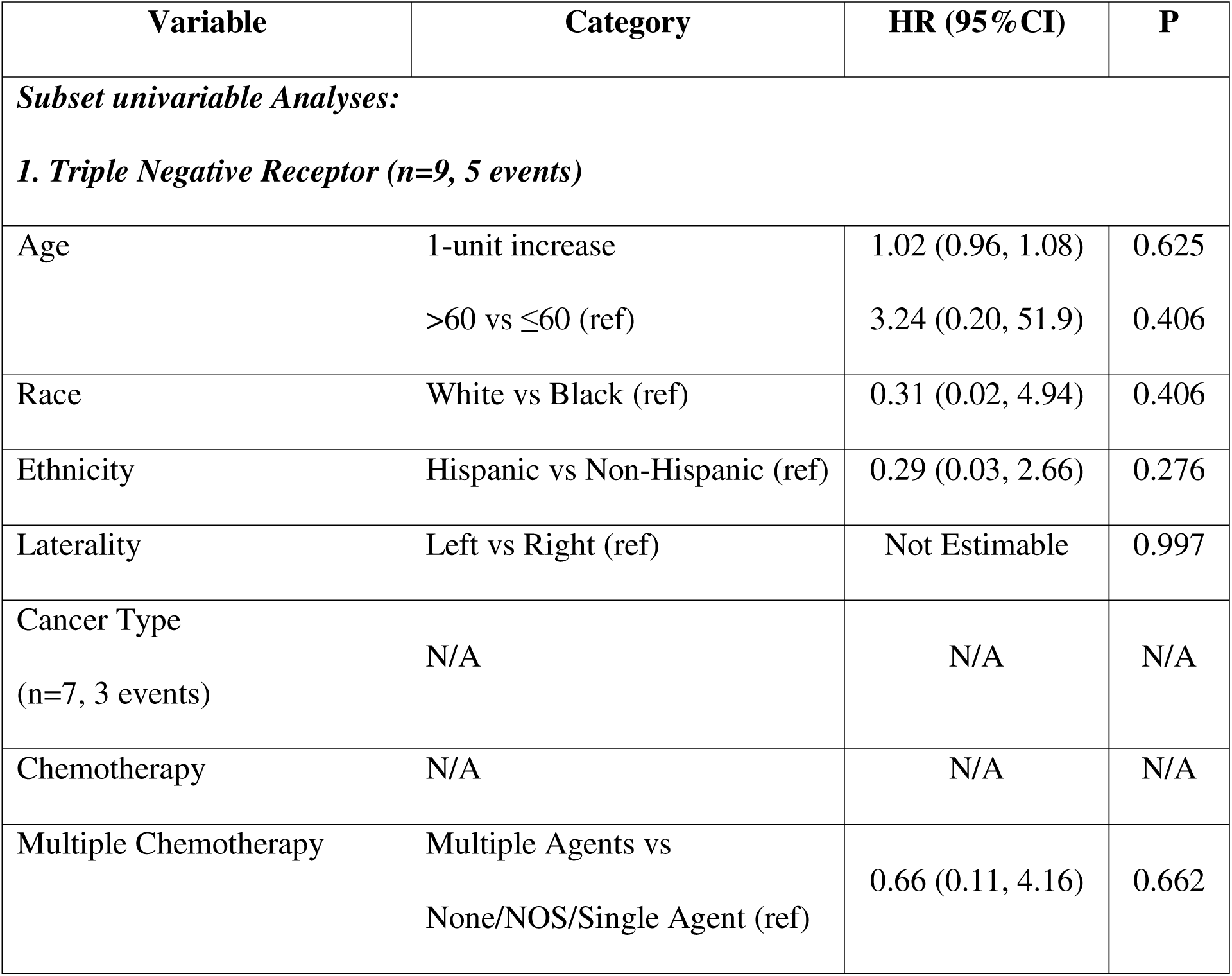
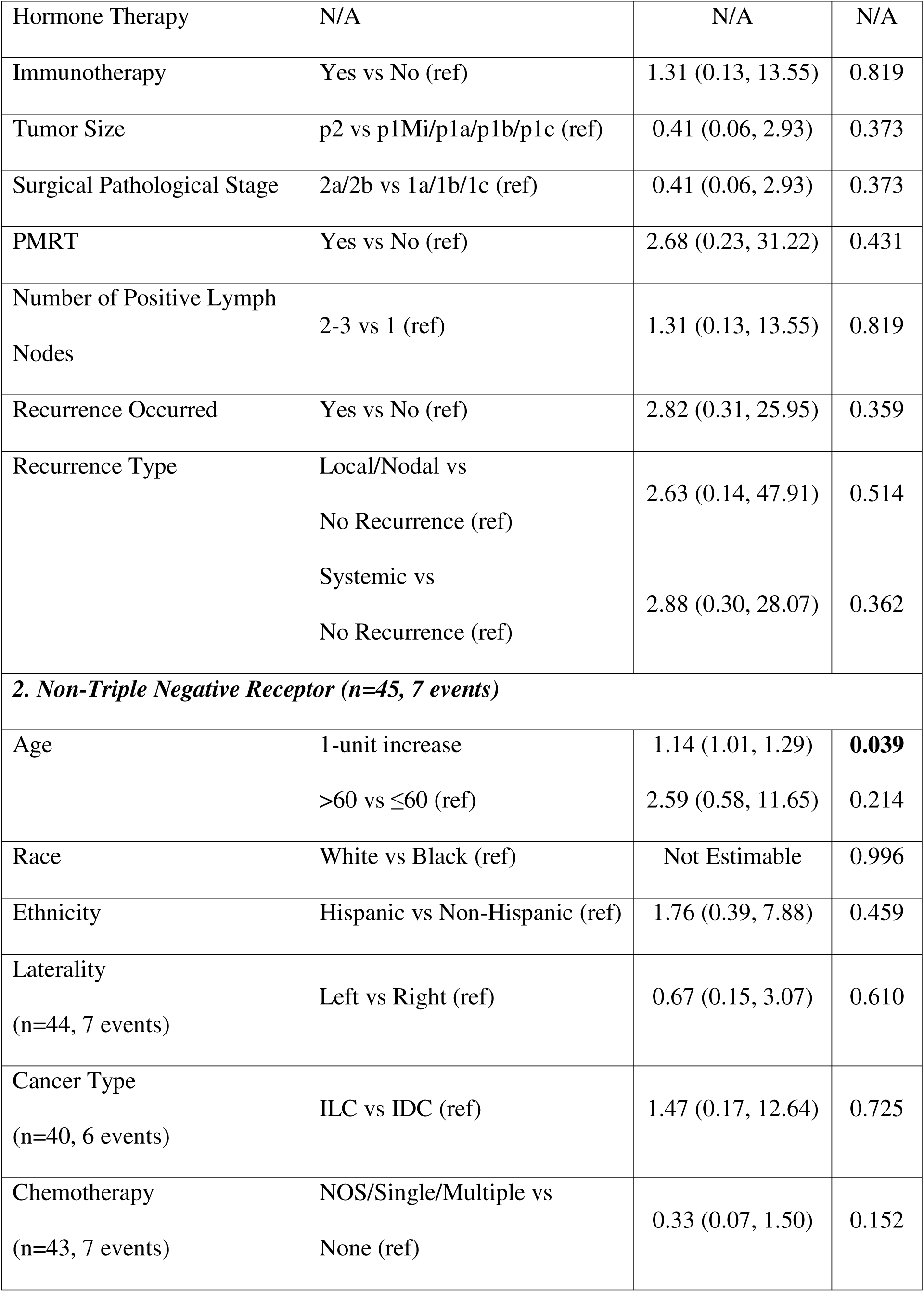
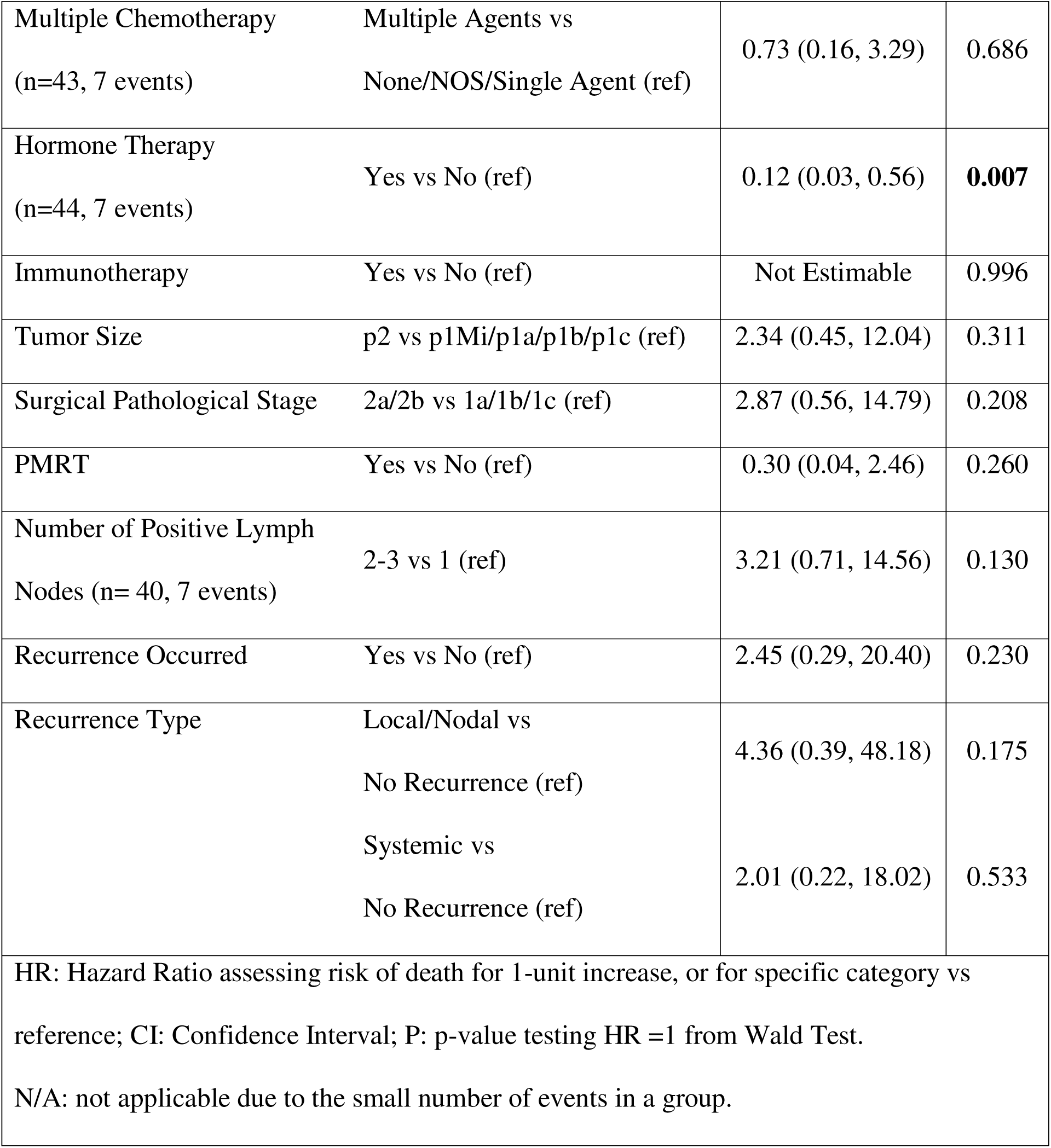
Cox Regression Analyses Assessing Risk Factors for OS by Receptor Status (Triple negative vs. Non-triple negative)

## Discussion

Our study demonstrates no statistically significant difference in OS and RFS when comparing PMRT and non-PMRT groups with pN1 breast cancer, suggesting that PMRT is not necessary for pN1 breast cancer patients. Many studies have not shown improvement in OS in N1 breast cancer patients who receive PMRT, similar to our findings [14–16].

Similar prior studies have shown that PMRT can reduce locoregional recurrence and improve RFS in N1 breast cancer patients when comparing certain subgroups. These subgroups include patients with increased tumor size, younger age, increased number of node involvement, and triple-negative subtype [6, 17–19]. Consistent with these claims, involvement of multiple nodes and triple negative disease was associated with decreased survival in our cohort.

The literature suggests that PMRT may confer a benefit in pN1 triple negative breast cancer (TNBC) patients given the higher risk of logoregional recurrence in these patients [20, 21]. However, exploratory subgroup analyses did not demonstrate a statistically significant differential benefit from PMRT by receptor subtype (Figure 3, Figure 4). These findings should be interpreted cautiously given the small number of triple-negative cases (n=9), particularly among patients receiving PMRT.

Notably, the absolute benefit of PMRT in N1 disease may be modest depending on response to systemic chemotherapy and other therapies. For instance, Wang et al. demonstrated a statistically significant improvement in survival parameters for all patients who received PMRT even after receiving systemic chemotherapy. However, for patients with pathologic complete response to systemic therapy, both RFS and OS did not improve after PMRT [22]. This further adds to the nuance and calls for an individualized approach to PMRT for pN1 breast cancer patients. Our study adds to the existing pool of literature to help characterize these cohorts.

Of note, stratification tools such as the 21-gene recurrence score (RS) assay have been studied to identify patients who may gain survival benefit from PMRT. A large observational cohort study of T1-2 N1 estrogen receptor-positive breast cancer patients from the National Cancer Database (NCDB) showed that patients with a low RS score had improved overall survival with PMRT [23, 24]. Yet a later study by Zhang et al. showed no difference in overall survival regardless of RS [25]. Together with the present findings, these data highlight the heterogeneity of pN1 breast cancer and suggest that molecular and biologic features, including receptor status, may inform risk stratification, though definitive subtype-specific benefit from PMRT remains unproven. This contributes to our understanding of a “no one size fits all” model and advocates for an individualized treatment plan for N1 breast cancer patients.

On subgroup analysis, patients who did not receive PMRT with left-sided tumor laterality had improved RFS and OS compared to those with right-sided tumors. The decreased prognosis in left-sided breast cancer compared to right-sided disease is consistent with outside literature [26]. Improved survival without radiation is also expected, as radiation to the left chest wall may cause long-term damage to nearby structures of the heart and lungs. Radiation therapy in left-sided breast cancer is associated with coronary artery disease, increased 10-year risk of myocardial infarctions, radiation-induced pneumonitis, and other related conditions [8, 27–29]. All of these secondary effects worsen mortality. Therefore, this study’s finding parallels external findings. This study is limited by its retrospective design, which introduces selection bias and limits the ability to establish causal relationships. There was variability in systemic therapy regimens including chemotherapy, endocrine therapy, and targeted agents as well as differences in surgical techniques across treating physicians and over time, which may have influenced outcomes. Pathologic and molecular data were occasionally incomplete, with some patients missing HER2 status, Ki67, or other prognostic biomarkers, which could limit risk stratification. Subgroup analyses stratified by receptor subtype were limited by small sample size and event counts and should be considered exploratory. Follow-up duration was heterogeneous and may not be sufficient to capture late toxicities, second malignancies, or long-term survival differences. Residual confounding from unmeasured variables such as comorbidities, adherence to adjuvant systemic therapy, and socioeconomic factors may have impacted both treatment selection and outcomes. This study also did not collect toxicity or quality of life data, which prevents evaluation of the morbidity associated with postmastectomy radiation therapy such as lymphedema, cardiopulmonary toxicity, and other late effects. These limitations highlight the need for larger prospective multi-institutional studies with standardized treatment protocols, comprehensive molecular profiling, and inclusion of patient-reported outcomes to better define the role of postmastectomy radiation therapy in node positive breast cancer.

## Conclusion

There remains a need for prospective, randomized trials to clarify the role of PMRT in pN1 breast cancer in the context of modern systemic therapy. Existing evidence is largely retrospective or derived from subgroup analyses, with heterogeneous patient populations and sometimes conflicting results. In our single-institution retrospective cohort, PMRT was not associated with statistically significant differences in RFS or OS; however, these findings should be interpreted cautiously given the modest sample size and limited power to detect modest effect. These findings underscore the heterogeneity of pN1 breast cancer and support continued efforts toward personalized, risk-adapted PMRT selection, ideally validated in larger multi-institutional prospective cohorts incorporating contemporary systemic therapy and tumor biology.

## Data Availability Statement

Data are stored in a secure institutional repository and may be accessed by qualified researchers upon reasonable request to the corresponding author.

## Acknowledgements

None

## References

[1] M. Overgaard et al., “Postoperative radiotherapy in high-risk premenopausal women with breast cancer who receive adjuvant chemotherapy. Danish Breast Cancer Cooperative Group 82b Trial,” N Engl J Med, vol. 337, no. 14, pp. 949–55, Oct 2 1997, doi: 10.1056/NEJM199710023371401.

[2] M. Overgaard et al., “Postoperative radiotherapy in high-risk postmenopausal breast-cancer patients given adjuvant tamoxifen: Danish Breast Cancer Cooperative Group DBCG 82c randomised trial,” (in eng), Lancet, vol. 353, no. 9165, pp. 1641–8, May 15 1999, doi: 10.1016/s0140-6736(98)09201-0.

[3] P. McGale et al., “Effect of radiotherapy after mastectomy and axillary surgery on 10-year recurrence and 20-year breast cancer mortality: meta-analysis of individual patient data for 8135 women in 22 randomised trials,” (in eng), Lancet, vol. 383, no. 9935, pp. 2127–35, Jun 21 2014, doi: 10.1016/s0140-6736(14)60488-8.

[4] W. J. Gradishar et al., “NCCN Guidelines® Insights: Breast Cancer, Version 4.2023,” (in eng), J Natl Compr Canc Netw, vol. 21, no. 6, pp. 594–608, Jun 2023, doi: 10.6004/jnccn.2023.0031.

[5] S.-F. Lai, C.-S. Huang, and S.-H. Kuo, “Whether adjuvant radiotherapy is desired for postmastectomy patients with T1–T2 tumors and 1–3 positive axillary lymph nodes who received modern systemic therapy?,” Translational Cancer Research, pp. S110–S114, 2018.

[6] P. T. Truong, I. A. Olivotto, H. A. Kader, M. Panades, C. H. Speers, and E. Berthelet, “Selecting breast cancer patients with T1-T2 tumors and one to three positive axillary nodes at high postmastectomy locoregional recurrence risk for adjuvant radiotherapy,” Int J Radiat Oncol Biol Phys, vol. 61, no. 5, pp. 1337–47, Apr 1 2005, doi: 10.1016/j.ijrobp.2004.08.009.

[7] J. Wei, Y. Jiang, and Z. Shao, “The survival benefit of postmastectomy radiotherapy for breast cancer patients with T1-2N1 disease according to molecular subtype,” The Breast, vol. 51, pp. 40–49, 2020/06/01/ 2020, doi: 10.1016/j.breast.2020.03.003.

[8] S. C. Darby et al., “Risk of ischemic heart disease in women after radiotherapy for breast cancer,” N Engl J Med, vol. 368, no. 11, pp. 987–98, Mar 14 2013, doi: 10.1056/NEJMoa1209825.

[9] A. Recht et al., “Postmastectomy Radiotherapy: An American Society of Clinical Oncology, American Society for Radiation Oncology, and Society of Surgical Oncology Focused Guideline Update,” J Clin Oncol, vol. 34, no. 36, pp. 4431–4442, Dec 20 2016, doi: 10.1200/JCO.2016.69.1188.

[10] I. Kunkler, “No OS benefit at 10 years with post mastectomy RT in intermediate risk breast cancer: 10 year overall survival results of the SUPREMO (BIG 2 04 MRC) randomized trial,” in SABCS 2024 Specialisation: Oncology, San Antonio, TX, USA, 2024/12 2024: Medicom Medical Publishers, doi: 10.55788/7455c8e0. [Online]. Available: https://conferences.medicom-publishers.com/specialisation/oncology/sabcs-2024/no-os-benefit-at-10-years-with-post-mastectomy-rt-in-intermediate-risk-breast-cancer-2/

[11] A. L. V. Johansson, C. B. Trewin, I. Fredriksson, K. V. Reinertsen, H. Russnes, and G. Ursin, “In modern times, how important are breast cancer stage, grade and receptor subtype for survival: a population-based cohort study,” Breast Cancer Research, vol. 23, no. 1, p. 17, 2021/02/01 2021, doi: 10.1186/s13058-021-01393-z.

[12] I. Soerjomataram, M. W. Louwman, J. G. Ribot, J. A. Roukema, and J. W. Coebergh, “An overview of prognostic factors for long-term survivors of breast cancer,” (in eng), Breast Cancer Res Treat, vol. 107, no. 3, pp. 309–30, Feb 2008, doi: 10.1007/s10549-007-9556-1.

[13] B. S. Jang and K. H. Shin, “Postmastectomy Radiation Therapy in Patients With Minimally Involved Lymph Nodes: A Review of the Current Data and Future Directions,” (in eng), J Breast Cancer, vol. 25, no. 1, pp. 1–12, Feb 2022, doi: 10.4048/jbc.2022.25.e6.

[14] H. Yin et al., “Impact of postmastectomy radiation therapy in T1-2 breast cancer patients with 1-3 positive axillary lymph nodes,” Oncotarget, vol. 8, no. 30, pp. 49564–49573, Jul 25 2017, doi: 10.18632/oncotarget.17318.

[15] F. Y. Li et al., “Real-world impact of postmastectomy radiotherapy in T1-2 breast cancer with one to three positive lymph nodes,” Ann Transl Med, vol. 8, no. 7, p. 489, Apr 2020, doi: 10.21037/atm.2020.03.49.

[16] M. Luo et al., “Postmastectomy radiotherapy in patients with T(1-2)N(1) breast cancer: a single center experience and a meta-analysis,” J Cancer Res Clin Oncol, vol. 149, no. 12, pp. 9979–9990, Sep 2023, doi: 10.1007/s00432-023-04908-7.

[17] A. Yamada et al., “Prognostic impact of postoperative radiotherapy in patients with breast cancer and with pT1-2 and 1-3 lymph node metastases: A retrospective cohort study based on the Japanese Breast Cancer Registry,” (in eng), Eur J Cancer, vol. 172, pp. 31–40, Sep 2022, doi: 10.1016/j.ejca.2022.05.017.

[18] D. Huo, N. Hou, N. Jaskowiak, D. J. Winchester, D. P. Winchester, and K. Yao, “Use of Postmastectomy Radiotherapy and Survival Rates for Breast Cancer Patients with T1-T2 and One to Three Positive Lymph Nodes,” Ann Surg Oncol, vol. 22, no. 13, pp. 4295–304, Dec 2015, doi: 10.1245/s10434-015-4528-x.

[19] E. Jwa et al., “Identification of Risk Factors for Locoregional Recurrence in Breast Cancer Patients with Nodal Stage N0 and N1: Who Could Benefit from Post-Mastectomy Radiotherapy?,” (in eng), PLoS One, vol. 10, no. 12, p. e0145463, 2015, doi: 10.1371/journal.pone.0145463.

[20] X. Chen et al., “Radiotherapy can improve the disease-free survival rate in triple-negative breast cancer patients with T1-T2 disease and one to three positive lymph nodes after mastectomy,” (in eng), Oncologist, vol. 18, no. 2, pp. 141–7, 2013, doi: 10.1634/theoncologist.2012-0233.

[21] H. J. Park et al., “Possible benefits from post-mastectomy radiotherapy in node-negative breast cancer patients: a multicenter analysis in Korea (KROG 14-22),” (in eng), Oncotarget, vol. 8, no. 35, pp. 59800–59809, Aug 29 2017, doi: 10.18632/oncotarget.16241.

[22] Q. Wang et al., “Is There a Role for Post-Mastectomy Radiotherapy for T1-2N1 Breast Cancers With Node-Positive Pathology After Patients Become Node-Negative Pathology Following Neoadjuvant Chemotherapy?,” (in eng), Front Oncol, vol. 10, p. 892, 2020, doi: 10.3389/fonc.2020.00892.

[23] C. R. Goodman, B. L. Seagle, M. Kocherginsky, E. D. Donnelly, S. Shahabi, and J. B. Strauss, “21-Gene Recurrence Score Assay Predicts Benefit of Post-Mastectomy Radiotherapy in T1-2 N1 Breast Cancer,” Clin Cancer Res, vol. 24, no. 16, pp. 3878–3887, Aug 15 2018, doi: 10.1158/1078-0432.CCR-17-3169.

[24] G. G. Halfteck, Y. E. Polychronopoulou, W. Haque, R. Weiser, S. S. Hatch, and V. S. Klimberg, “De-escalation of Post-mastectomy Irradiation in Hormone Receptor-Positive Breast Cancer with One to Three Positive Nodes,” Ann Surg Oncol, vol. 30, no. 13, pp. 8335–8343, Dec 2023, doi: 10.1245/s10434-023-14155-2.

[25] W. W. Zhang et al., “21-Gene Recurrence Score Assay Could Not Predict Benefit of Post-mastectomy Radiotherapy in T1-2 N1mic ER-Positive HER2-Negative Breast Cancer,” Front Oncol, vol. 9, p. 270, 2019, doi: 10.3389/fonc.2019.00270.

[26] Y. Abdou et al., “Left sided breast cancer is associated with aggressive biology and worse outcomes than right sided breast cancer,” Sci Rep, vol. 12, no. 1, p. 13377, Aug 4 2022, doi: 10.1038/s41598-022-16749-4.

[27] M. J. Hooning et al., “Long-term risk of cardiovascular disease in 10-year survivors of breast cancer,” J Natl Cancer Inst, vol. 99, no. 5, pp. 365–75, Mar 7 2007, doi: 10.1093/jnci/djk064.

[28] V. Mehta, “Radiation pneumonitis and pulmonary fibrosis in non-small-cell lung cancer: pulmonary function, prediction, and prevention,” Int J Radiat Oncol Biol Phys, vol. 63, no. 1, pp. 5–24, Sep 1 2005, doi: 10.1016/j.ijrobp.2005.03.047.

[29] I. Madani, K. De Ruyck, H. Goeminne, W. De Neve, H. Thierens, and J. Van Meerbeeck, “Predicting risk of radiation-induced lung injury,” J Thorac Oncol, vol. 2, no. 9, pp. 864–74, Sep 2007, doi: 10.1097/JTO.0b013e318145b2c6.

